# CMS Final Rule for Organ Donation: Unintended Consequences of the Pancreas Research Loophole

**DOI:** 10.1101/2023.03.19.23287457

**Authors:** David Goldberg, Darius Chyou, Rachael Wulf, Matthew Wadsworth

**Affiliations:** Division of Digestive Health and Liver Diseases, University of Miami Miller School of Medicine, Miami, FL; University of Miami/Jackson Memorial Hospital, Department of Medicine, Miami, Fl; Life Connection of Ohio, Toledo, OH

**Keywords:** donation, metrics, policy

## Abstract

CMS updated the ‘Final Rule’ for OPO Conditions for Coverage on 11/20/2020 to include new OPO metrics, tiers of classification, and a new definition of a donor that included an individual with: a) ≥1 organ transplanted; or b) pancreas procured for research or islet cell transplantation. We conducted a retrospective cohort study using data from the OPTN/ UNOS. The number of pancreata procured for research increased four-fold from 2020 to 2022, despite stable numbers of other organs procured for research. Of the 57 OPOs, 8 (14.0%) procured >100 pancreata for research in 2022, accounting for 1,548 (58.2%) pancreata research procurements. Of those 8 OPOs, seven would be at risk for possible (Tier 2; n=3) or definite (Tier 3; n=4) decertification based on CMS’s 2022 interim report. Based on these data, two Tier 3 OPOs would be reclassified as Tier 2 and one as Tier 1; three Tier 2 OPOs would be reclassified as Tier 1; and three Tier 3s would be reclassified as borderline Tier 2. The pancreas research carveout may detrimentally impact the system of organ donation by failing to accurately measure OPO performance and flag (and decertify) underpeforming OPOs unless CMS revises its final rule.

## INTRODUCTION

Organ procurement organizations (OPOs) are the federal contractors who manage all aspects of deceased organ donation in the United States. The issue of variability in OPO performance received attention from the highest levels of government in 2019, leading to The Advancing American Kidney Health Initiative Executive Order in 2019. As a result of this executive order, the Centers for Medicare and Medicaid Services (CMS) updated the ‘Final Rule’ for OPO Conditions for Coverage on 11/20/2020. This updated final rule redefined how OPOs would be evaluated using objective, standardized, verifiable data, with the metrics denominator relying on the cause-age-location-consistent with death (CALC) metric that utilized National Center for Health Statistics Death data.^1-3^ In addition to codifying new metrics (donation rates and transplantation rate), the final rule set criteria to classify OPOs into three tiers based on the confidence intervals around their donation and transplantation metrics. At the end of each four-year certification cycle, Tier 1 OPOs (i.e., the highest-performing OPOs) would be re-certified, Tier 2 OPOs would be at risk for potential decertification (or competition to retain their donation service area), while Tier 3 OPOs would be decertified.^2^ As part of the final rule, CMS redefined a donor for regulatory purposes, defined as an individual with: a) ≥1 organ transplanted; or b) pancreas procured for research or islet cell transplantation (only performed under research protocol).^1-3^ As a result of this pancreatic research carveout, a Tier 3 OPO could procure more pancreata for research to avoid definite decertification, and a Tier 2 OPO could avoid potential decertification (or competition to retain their donation service area) without facilitating more transplants.^2^ We sought to evaluate the impact of the final rule on procurement of research pancreata and how it might change the tier classification of an OPO.

## METHODS

We conducted a retrospective cohort study using data from the Organ Procurement and Transplantation Network (OPTN)/United Network for Organ Sharing (UNOS). We evaluated data over a 10-year period from 1/1/2013-12/31/2022. We used the deceased donor dataset that included data on all organ donors, and the disposition of each organ. Pancreata donated for research were identified based on OPTN/UNOS codes, and pancreas-only donors were those for whom a pancreas was donated for research, and no other organs were donated. To assess the impact of increased procurement of pancreata for research, we evaluated CMS’s 2022 OPO interim performance report to reclassify OPOs based on their increased volume of pancreata research procurement.^1,4^ In this report, it lists the number of donors and transplants each OPO would need to meet the 50^th^ percentile and 25^th^ percentile. Based on the pancreata procured for research and the tier classifications, Tier 2 and 3 OPOs were potentially reclassified if their increased pancreata (organs) and donors would meet the 50^th^ and 25^th^ percentile. This study was considered exempt by the Institutional Review Board at the University of Miami.

## RESULTS

Despite a steady increase in donors with ≥1 organ transplanted, there was a striking increase in individuals classified as a donor solely because their pancreas was procured for research (Figure 1a). The number of pancreata procured for research increased four-fold from 2020 to 2022, despite stable numbers of other organs procured for research (Figure 1b), with a progressive monthly increase in pancreata procured for research (Figure 1c).

**Figure 1.**
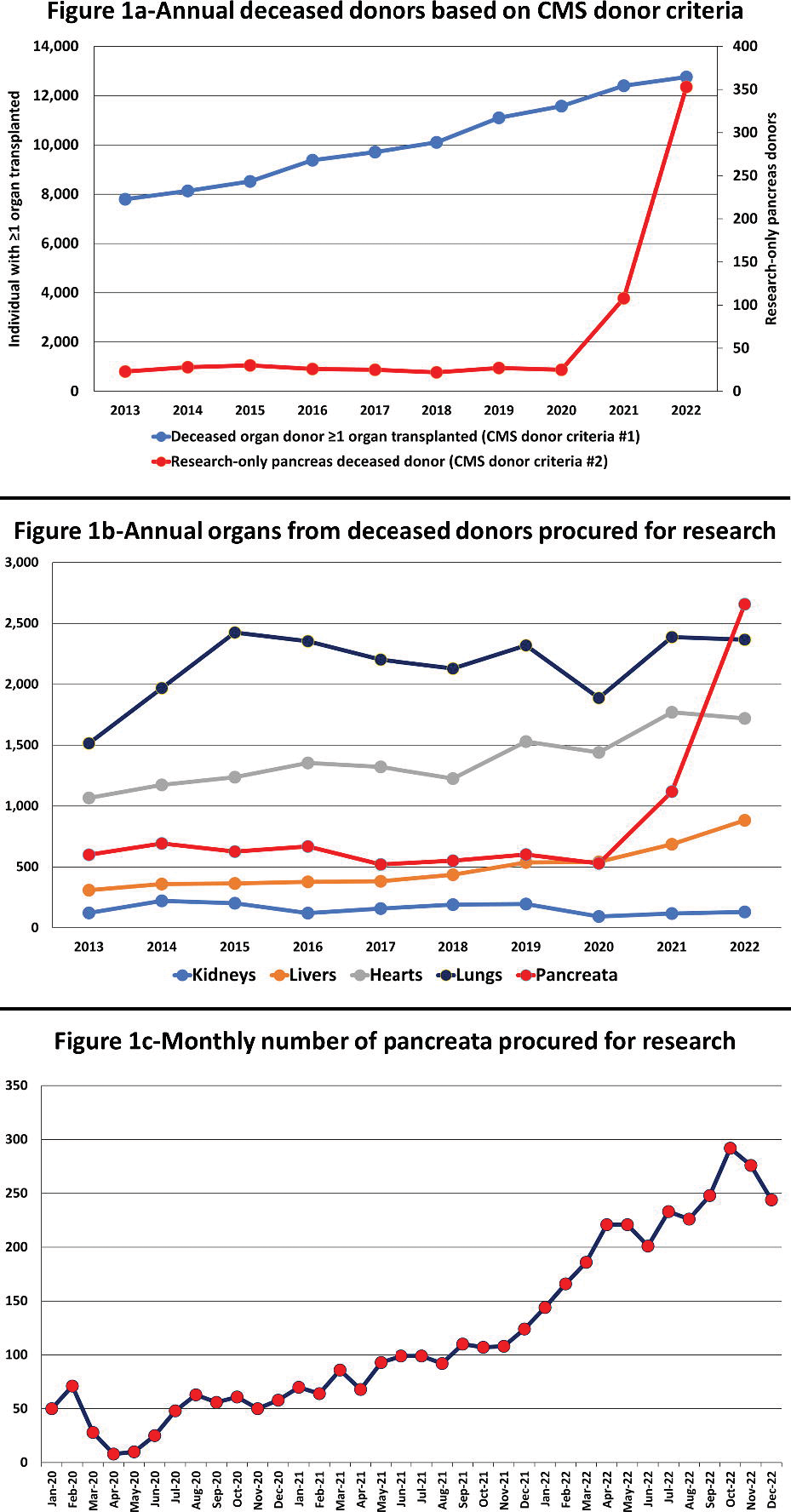
(three panels): National changes in procurement of pancreata for research a. Figure 1a: Annual number of deceased donors based on CMS criteria of ≥1 organ transplant or a pancreas procured for research or islet cells from 2013-2022 b. Figure 1b: Annual number of organs procured from deceased donors for research purposes from 2013-2022 c. Figure 1c: Monthly number of pancreata procured for research from 2020-2022

The increase in pancreata procured for research varied across OPOs (Figures 2a, 2b). Of the 57 OPOs, 8 (14.0%) procured >100 pancreata for research in 2022, accounting for 1,548 (58.2%) pancreata research procurements. Of those 8 OPOs, seven would be at risk for possible (Tier 2; n=3) or definite (Tier 3; n=4) decertification based on CMS’s 2022 interim report.^1,4^ Importantly, based on these data, two Tier 3 OPOs would be reclassified as Tier 2 (Louisiana Organ Procurement Agency, LAOP; LifeBanc, OHLB) and one as Tier 1 (LifeCenter Organ Donor Network; OHOV); three Tier 2 OPOs would be reclassified as Tier 1 (Donor Network West, CADN; Donor Alliance, CORS; and Lifeline of Ohio, OHLP); and three Tier 3s would be reclassified as borderline Tier 2 (OneLegacy, CAOP); LifeQuest Organ Recovery Services, FLUF; and Kentucky Organ Donor Affiliates, KYDA).

**Figure 2.**
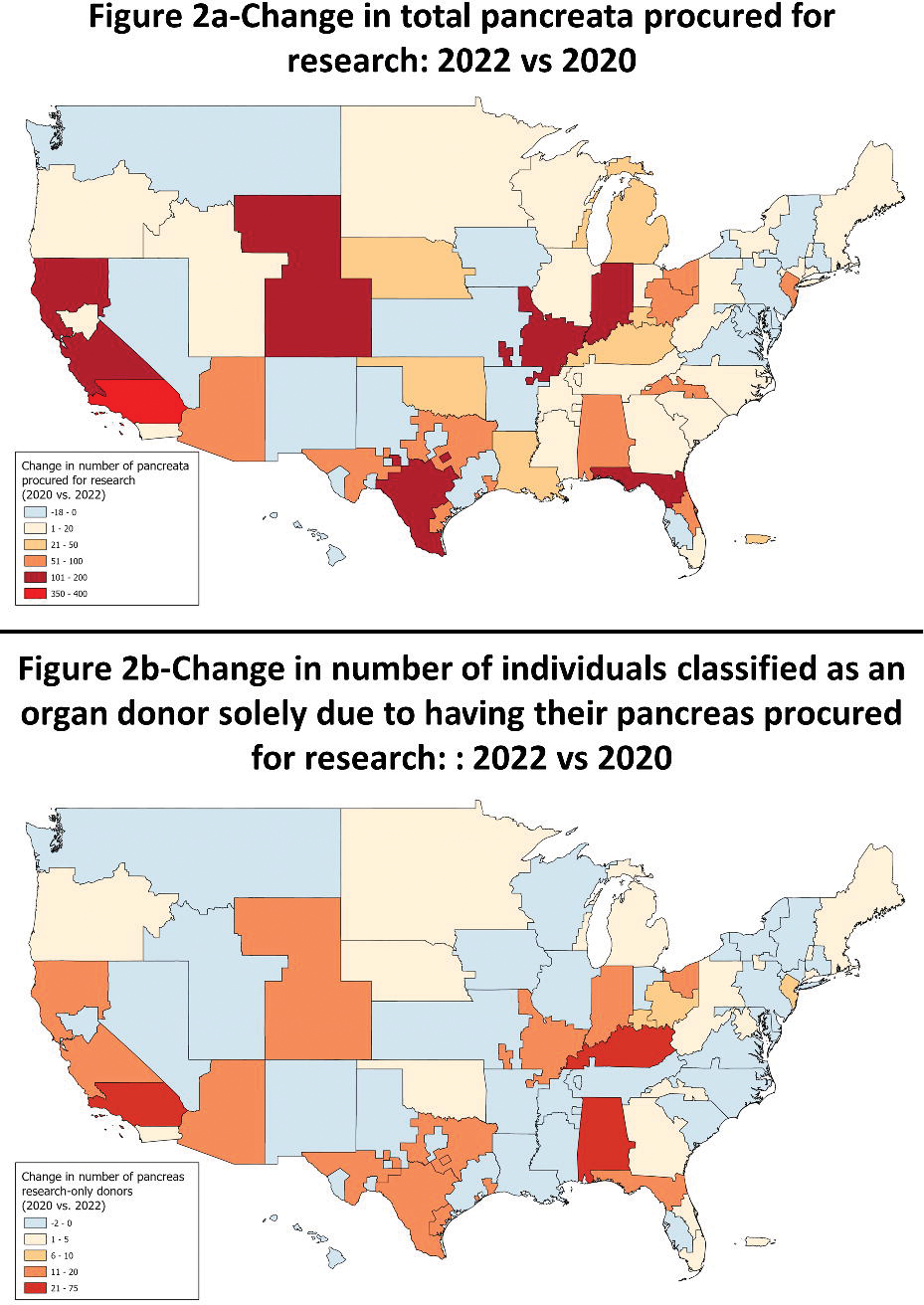
(two panels): OPO-level changes in procurement of pancreata for research in 2022 versus 202 a. Figure 2a: Change in total pancreata procured for research b. Figure 2b: Change in the number of individuals classified as an organ donor solely due to having their pancreas procured for research

## DISCUSSION

CMS updated the final rule for OPO performance to increase the number of transplants by better comparing OPOs using standardized objective data to identify low-performing OPOs that would/could face decertification.^1-3^ At the time, the pancreas research procurement carveout appeared misaligned with these goals.^1,5^ Our data reveal how the rule change led to an increase in pancreata procured for research, thereby improving the measured “performance” of several OPOs without translating to an increase in transplants (islet cell transplants are rare^5^). This would have led to three Tier 3 OPOs avoiding definite decertification, three Tier 2 OPOs avoiding potential decertification, and three Tier 3 OPOs being on the cusp of avoiding definite decertification.

We could not quantify which pancreata were used for research and/or islet cells, however the temporal relationship at select OPOs, nearly all of which were classified by CMS as underpeforming, suggest practice shifted due to the rule change. The pancreas research carevout is an unintended and negative consequence of a CMS rule change that improves the perceived performance of several underperforming OPOs, without yielding more transplants. This will detrimentally impact the system of organ donation by failing to accurately measure OPO performance and flag (and decertify) underpeforming OPOs unless CMS revises its final rule.

## Data Availability

All data produced in the present work are contained in the manuscript and the source code can be shared upon request.

## Acknowledgments

This work was supported in part by Health Resources and Services Administration contract HHSH250-2019-00001C. The content is the responsibility of the authors alone and does not necessarily reflect the views or policies of the Department of Health and Human Services, nor does mention of trade names, commercial products, or organizations imply endorsement by the U.S. Government.

